# Clinical coding of long COVID in English primary care: a federated analysis of 58 million patient records in situ using OpenSAFELY

**DOI:** 10.1101/2021.05.06.21256755

**Authors:** The OpenSAFELY Collaborative, Alex J Walker, Brian MacKenna, Peter Inglesby, Christopher T Rentsch, Helen J Curtis, Caroline E Morton, Jessica Morley, Amir Mehrkar, Seb Bacon, George Hickman, Chris Bates, Richard Croker, David Evans, Tom Ward, Jonathan Cockburn, Simon Davy, Krishnan Bhaskaran, Anna Schultze, Elizabeth J Williamson, William J Hulme, Helen I McDonald, Laurie Tomlinson, Rohini Mathur, Rosalind M Eggo, Kevin Wing, Angel YS Wong, Harriet Forbes, John Tazare, John Parry, Frank Hester, Sam Harper, Shaun O’Hanlon, Alex Eavis, Richard Jarvis, Dima Avramov, Paul Griffiths, Aaron Fowles, Nasreen Parkes, Ian J Douglas, Stephen JW Evans, Liam Smeeth, Ben Goldacre

## Abstract

**Background:** Long COVID is a term to describe new or persistent symptoms at least four weeks after onset of acute COVID-19. Clinical codes to describe this phenomenon were released in November 2020 in the UK, but it is not known how these codes have been used in practice.

**Methods:** Working on behalf of NHS England, we used OpenSAFELY data encompassing 96% of the English population. We measured the proportion of people with a recorded code for long COVID, overall and by demographic factors, electronic health record software system, and week. We also measured variation in recording amongst practices.

**Results:** Long COVID was recorded for 23,273 people. Coding was unevenly distributed amongst practices, with 26.7% of practices having not used the codes at all. Regional variation was high, ranging between 20.3 per 100,000 people for East of England (95% confidence interval 19.3-21.4) and 55.6 in London (95% CI 54.1-57.1). The rate was higher amongst women (52.1, 95% CI 51.3-52.9) compared to men (28.1, 95% CI 27.5-28.7), and higher amongst practices using EMIS software (53.7, 95% CI 52.9-54.4) compared to TPP software (20.9, 95% CI 20.3-21.4).

**Conclusions:** Long COVID coding in primary care is low compared with early reports of long COVID prevalence. This may reflect under-coding, sub-optimal communication of clinical terms, under-diagnosis, a true low prevalence of long COVID diagnosed by clinicians, or a combination of factors. We recommend increased awareness of diagnostic codes, to facilitate research and planning of services; and surveys of clinicians’ experiences, to complement ongoing patient surveys.

## Background

Long COVID has been broadly defined as new or persistent symptoms of COVID-19 beyond the acute phase of SARS-CoV-2 infection. The National Institute for Health and Care Excellence (NICE) have produced guidance on managing the long-term effects of COVID-19 as these symptoms can have a significant effect on a person’s quality of life.^1^ NICE recognise that as long COVID is such a new condition the exact clinical definition and treatments are evolving.

NICE developed their definitions and clinical guidelines using a “living” approach based on early data. This means that the guidelines will be continuously reviewed and updated and it is therefore critical to continue studying the long-term effects of COVID-19 as data accrue, and refine the guidelines appropriately. To support this need, long COVID SNOMED-CT codes (the “diagnostic codes” in Table 3) were developed and released in the UK in November 2020. To support clinical care and implementation of NICE guidance, distinct SNOMED-CT codes are available which distinguish between the length of ongoing symptoms. Symptoms between 4-12 weeks are defined as “ongoing symptomatic COVID-19”, and symptoms continuing beyond 12 weeks as “post-COVID-19 syndrome”.^2^ There are also 3 assessment codes and 10 referral codes relating to long COVID.

We set out to describe the use of long COVID codes in English primary care since their introduction, in a cohort of covering approximately 96% of the English population - those covered by the two largest electronic health record providers, EMIS and TPP (SystmOne). We describe the distribution of use amongst general practices, demographics and over time.

## Methods

### Study design and data sources

We conducted a study calculating the period prevalence of long COVID recording in electronic health record (EHR) data. Primary care records managed by the GP software providers EMIS and TPP were accessed through OpenSAFELY, an open source data analytics platform created by our team on behalf of NHS England to address urgent COVID-19 research questions (https://opensafely.org). OpenSAFELY provides a secure software interface allowing a federated analysis of pseudonymized primary care patient records from England in near real-time within the EMIS and TPP highly secure data environments. Non-disclosive, aggregated results are exported to GitHub where further data processing and analysis takes place. This avoids the need for large volumes of potentially disclosive pseudonymised patient data to be transferred off-site. This, in addition to other technical and organisational controls, minimizes any risk of re-identification. The dataset available to the platform includes pseudonymised data such as coded diagnoses, medications and physiological parameters. No free text data are included. All activity on the platform is publicly logged and all analytic code and supporting clinical coding lists are automatically published. In addition, the framework provides assurance that the analysis is reproducible and reusable. Further details on our information governance and platform can be found in the Appendix under information governance and ethics.

### Population

We included all people registered with a general practice on the 1st November 2020.

### Outcome

The outcome was any record of long COVID in the primary care record. This was defined using a list of 15 UK SNOMED codes, which are listed in Table 3 and categorised as diagnostic (2 codes), referral (3) and assessment (10).^3^ The outcome was measured between the study start date (2020-02-01) and the end date (2021-04-25). Though the start date is before the codes were created, it’s possible to backdate diagnostic codes in a GP system. Timing of outcomes was determined by the first record of a SNOMED code for each person, as determined by the date recorded by the clinician.

### Stratifiers

Demographic variables were extracted including age (in categories), sex, geographic region, Index of Multiple Deprivation (IMD, divided into quintiles), and ethnicity. Counts and rates of recorded events were stratified by each demographic variable. Recording of each SNOMED code was assessed individually, in this case counting every recorded code including repeated codes, rather than one per patient.

### Statistical methods

We calculated proportions of patients with long COVID codes over the whole study period per 100,000 patients, 95% confidence intervals of those proportions, and the distribution of codes by each stratification variable.

### Software and Reproducibility

Data management and analysis was performed using the OpenSAFELY software libraries and Jupyter notebooks, both implemented using Python 3. More details are available in the Appendix. This is an analysis delivered using federated analysis through the OpenSAFELY platform: codelists and code for data management and data analysis were specified once using the OpenSAFELY tools; then transmitted securely from the OpenSAFELY jobs server to the OpenSAFELY-TPP platform within TPP’s secure environment, and separately to the OpenSAFELY-EMIS platform within EMIS’s secure environment, where they were each executed separately against local patient data; summary results were then reviewed for disclosiveness, released, and combined for the final outputs. All code for the OpenSAFELY platform for data management, analysis and secure code execution is shared for review and re-use under open licenses at GitHub.com/OpenSAFELY. All code for data management and analysis for this paper is shared for scientific review and re-use under open licenses on GitHub https://github.com/opensafely/long-covid.

## Results

### Cohort characteristics and overall rate of recording

There were 58.0m people in the combined cohort in total, 24.0m in the TPP cohort, and 34.0m in the EMIS cohort. Demographics of the cohort are described in Table 1. Up to 25th April 2021, there were 23,273 patients with a recorded code indicative of long COVID diagnosis. A higher proportion of these recorded diagnoses were in EMIS, with 18,262, compared to 5,011 in TPP. Taking into account the larger total number of patients in EMIS practices, the rate over the whole study period was 53.7 per 100,000 people (95% CI 52.9-54.4) in EMIS and 20.9 (95% CI 20.3-21.4) in TPP.

**Table 1:** Characteristics of the cohort

| Attribute | Category | TPP |  | EMIS |  | Combined |  |
| --- | --- | --- | --- | --- | --- | --- | --- |
|  |  | Total | % | Total | % | Total | % |
| **Total** |  | 24,011,964 | 100.0 | 34,032,530 | 100.0 | 58,044,494 | 100.0 |
| **Age group** | 0-17 | 4,821,223 | 20.1 | 6,901,845 | 20.3 | 11,723,068 | 20.2 |
|  | 18-24 | 1,901,509 | 7.9 | 2,884,964 | 8.5 | 4,786,473 | 8.2 |
|  | 25-34 | 3,340,123 | 13.9 | 4,962,526 | 14.6 | 8,302,649 | 14.3 |
|  | 35-44 | 3,220,499 | 13.4 | 4,745,812 | 13.9 | 7,966,311 | 13.7 |
|  | 45-54 | 3,230,861 | 13.5 | 4,546,614 | 13.4 | 7,777,475 | 13.4 |
|  | 55-69 | 4,202,414 | 17.5 | 5,697,231 | 16.7 | 9,899,645 | 17.1 |
|  | 70-79 | 2,080,859 | 8.7 | 2,699,998 | 7.9 | 4,780,857 | 8.2 |
|  | 80+ | 1,214,476 | 5.1 | 1,593,540 | 4.7 | 2,808,016 | 4.8 |
| **Sex** | F | 12,004,974 | 50.0 | 17,014,169 | 50.0 | 29,019,143 | 50.0 |
|  | M | 12,006,990 | 50.0 | 17,018,361 | 50.0 | 29,025,351 | 50.0 |
| **Region** | East of England | 5,638,753 | 23.5 | 1,341,520 | 3.9 | 6,980,273 | 12.0 |
|  | East Midlands | 4,191,051 | 17.5 | 763,830 | 2.2 | 4,954,881 | 8.5 |
|  | London | 1,702,673 | 7.1 | 7,804,070 | 22.9 | 9,506,743 | 16.4 |
|  | North East | 1,100,356 | 4.6 | 1,189,619 | 3.5 | 2,289,975 | 3.9 |
|  | North West | 2,067,131 | 8.6 | 6,875,180 | 20.2 | 8,942,311 | 15.4 |
|  | South East | 1,582,440 | 6.6 | 7,191,261 | 21.1 | 8,773,701 | 15.1 |
|  | South West | 3,304,393 | 13.8 | 2,488,558 | 7.3 | 5,792,951 | 10.0 |
|  | West Midlands | 988,286 | 4.1 | 5,057,090 | 14.9 | 6,045,376 | 10.4 |
|  | Yorkshire and The Humber | 3,427,713 | 14.3 | 1,278,147 | 3.8 | 4,705,860 | 8.1 |
| **IMD** | 1 Most deprived | 4,818,642 | 20.1 | 7,015,392 | 20.6 | 11,834,034 | 20.4 |
|  | 2 | 4,707,307 | 19.6 | 7,244,664 | 21.3 | 11,951,971 | 20.6 |
|  | 3 | 4,941,725 | 20.6 | 6,633,133 | 19.5 | 11,574,858 | 19.9 |
|  | 4 | 4,655,595 | 19.4 | 6,401,478 | 18.8 | 11,057,073 | 19.0 |
|  | 5 Least deprived | 4,302,292 | 17.9 | 6,635,613 | 19.5 | 10,937,905 | 18.8 |
| **Ethnicity** | White | 14,573,038 | 60.7 | 17,677,690 | 51.9 | 32,250,728 | 55.6 |
|  | Mixed | 319,793 | 1.3 | 581,965 | 1.7 | 901,758 | 1.6 |
|  | South Asian | 1,500,012 | 6.2 | 2,489,843 | 7.3 | 3,989,855 | 6.9 |
|  | Black | 515,866 | 2.1 | 1,173,341 | 3.4 | 1,689,207 | 2.9 |
|  | Other | 476,065 | 2.0 | 754,993 | 2.2 | 1,231,058 | 2.1 |

### Rate of coding stratified by demographics

Counts and rates of long COVID coding stratified by demographic factors are presented in Table 2. For age, the incidence of long COVID recording rises to a peak in the 45-54 group, before declining again in older age groups. Women had a higher rate of recording than men (52.1 (95% CI 51.3-52.9) vs 28.1 (95% CI 27.5-28.7) per 100,000 people). Counts of long COVID recording by IMD and ethnicity are reported in Table 2.

**Table 2:** Counts and rates of long COVID coding stratified by demographic variable

| Attribute | Category | TPP |  |  | EMIS |  |  | Combined |  |  |  |  |
| --- | --- | --- | --- | --- | --- | --- | --- | --- | --- | --- | --- | --- |
|  |  | Long COVID | Column % | Rate per 100,000 | Long COVID | Column % | Rate per 100,000 | Long COVID | Column % | Rate per 100,000 | Lower 95% CI | Upper 95% CI |
| Total |  | 5,011 | 100.0 | 20.9 | 18,262 | 100.0 | 53.7 | 23,273 | 100.0 | 40.1 | 39.6 | 40.6 |
| Age group | 0-17 | 94 | 1.9 | 1.9 | 248 | 1.4 | 3.6 | 342 | 1.5 | 2.9 | 2.6 | 3.2 |
|  | 18-24 | 177 | 3.5 | 9.3 | 684 | 3.7 | 23.7 | 861 | 3.7 | 18.0 | 16.8 | 19.2 |
|  | 25-34 | 592 | 11.8 | 17.7 | 2,267 | 12.4 | 45.7 | 2,859 | 12.3 | 34.4 | 33.2 | 35.7 |
|  | 35-44 | 1,033 | 20.6 | 32.1 | 4,077 | 22.3 | 85.9 | 5,110 | 22.0 | 64.1 | 62.4 | 65.9 |
|  | 45-54 | 1,392 | 27.8 | 43.1 | 5,183 | 28.4 | 114.0 | 6,575 | 28.3 | 84.5 | 82.5 | 86.6 |
|  | 55-69 | 1,361 | 27.2 | 32.4 | 4,869 | 26.7 | 85.5 | 6,230 | 26.8 | 62.9 | 61.4 | 64.5 |
|  | 70-79 | 261 | 5.2 | 12.5 | 693 | 3.8 | 25.7 | 954 | 4.1 | 20.0 | 18.7 | 21.2 |
|  | 80+ | 101 | 2.0 | 8.3 | 241 | 1.3 | 15.1 | 342 | 1.5 | 12.2 | 10.9 | 13.5 |
| Sex | F | 3,227 | 64.4 | 26.9 | 11,893 | 65.1 | 69.9 | 15,120 | 65.0 | 52.1 | 51.3 | 52.9 |
|  | M | 1,784 | 35.6 | 14.9 | 6,369 | 34.9 | 37.4 | 8,153 | 35.0 | 28.1 | 27.5 | 28.7 |
| Region | East of England | 913 | 18.2 | 16.2 | 505 | 2.8 | 37.6 | 1,418 | 6.1 | 20.3 | 19.3 | 21.4 |
|  | East Midlands | 775 | 15.5 | 18.5 | 314 | 1.7 | 41.1 | 1,089 | 4.7 | 22.0 | 20.7 | 23.3 |
|  | London | 265 | 5.3 | 15.6 | 5,021 | 27.5 | 64.3 | 5,286 | 22.7 | 55.6 | 54.1 | 57.1 |
|  | North East | 328 | 6.5 | 29.8 | 628 | 3.4 | 52.8 | 956 | 4.1 | 41.7 | 39.1 | 44.4 |
|  | North West | 395 | 7.9 | 19.1 | 4,185 | 22.9 | 60.9 | 4,580 | 19.7 | 51.2 | 49.7 | 52.7 |
|  | South East | 593 | 11.8 | 37.5 | 3,463 | 19.0 | 48.2 | 4,056 | 17.4 | 46.2 | 44.8 | 47.7 |
|  | South West | 797 | 15.9 | 24.1 | 1,004 | 5.5 | 40.3 | 1,801 | 7.7 | 31.1 | 29.7 | 32.5 |
|  | West Midlands | 288 | 5.7 | 29.1 | 2,598 | 14.2 | 51.4 | 2,886 | 12.4 | 47.7 | 46.0 | 49.5 |
|  | Yorkshire and The Humber | 655 | 13.1 | 19.1 | 528 | 2.9 | 41.3 | 1,183 | 5.1 | 25.1 | 23.7 | 26.6 |
| IMD | 1 Most deprived | 912 | 18.2 | 18.9 | 4,031 | 22.1 | 57.5 | 4,943 | 21.2 | 41.8 | 40.6 | 42.9 |
|  | 2 | 970 | 19.4 | 20.6 | 4,383 | 24.0 | 60.5 | 5,353 | 23.0 | 44.8 | 43.6 | 46.0 |
|  | 3 | 1,049 | 20.9 | 21.2 | 3,486 | 19.1 | 52.6 | 4,535 | 19.5 | 39.2 | 38.0 | 40.3 |
|  | 4 | 1,013 | 20.2 | 21.8 | 3,287 | 18.0 | 51.3 | 4,300 | 18.5 | 38.9 | 37.7 | 40.1 |
|  | 5 Least deprived | 949 | 18.9 | 22.1 | 3,034 | 16.6 | 45.7 | 3,983 | 17.1 | 36.4 | 35.3 | 37.5 |
| Ethnicity | White | 3,393 | 84.8 | 23.3 | 7,350 | 74.4 | 41.6 | 10,743 | 46.2 | 33.3 | 32.7 | 33.9 |
|  | Mixed | 63 | 1.6 | 19.7 | 223 | 2.3 | 38.3 | 286 | 1.2 | 31.7 | 28.0 | 35.4 |
|  | South Asian | 392 | 9.8 | 26.1 | 1,549 | 15.7 | 62.2 | 1,941 | 8.3 | 48.6 | 46.5 | 50.8 |
|  | Black | 91 | 2.3 | 17.6 | 560 | 5.7 | 47.7 | 651 | 2.8 | 38.5 | 35.6 | 41.5 |
|  | Other | 63 | 1.6 | 13.2 | 193 | 2.0 | 25.6 | 256 | 1.1 | 20.8 | 18.2 | 23.3 |

### Geographic and practice distribution of coding

The rate of coding varies substantially between regions (Table 2), from a minimum proportion of 20.3 per 100,000 people in the East of England (95%CI 19.3-21.4) to 55.6 in London (95% CI 54.1-57.1), though these region specific data were only available in TPP practices at the time of data extraction. Over a quarter (26.7%) of practices have not used the codes at all. This proportion is much higher in practices using TPP software (44.2%) than those using EMIS (15.1%). The distribution is described more fully in Figure 1. The highest number of codes in a single practice was 150.

**Figure 1:**
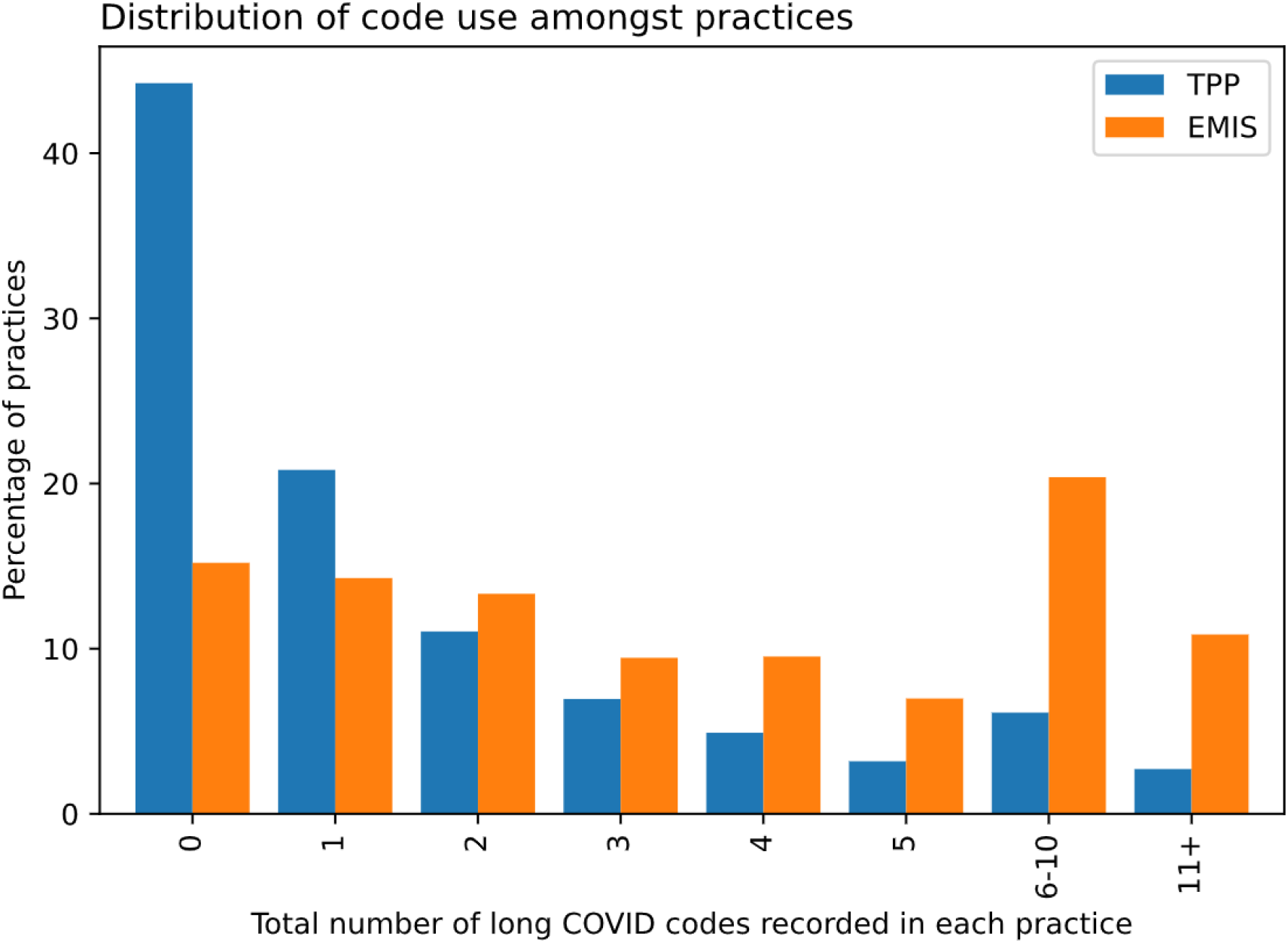
Volume of code use in individual practices, stratified by the electronic health record provider of the practice (TPP/SystmOne or EMIS).

### Rate of coding over time

The number of recorded events was relatively low until the end of January 2021, after which there was an increase in coding (Figure 2). This increase was more marked in EMIS practices, which before that time had recorded fewer long COVID codes overall than TPP practices.

**Figure 2:**
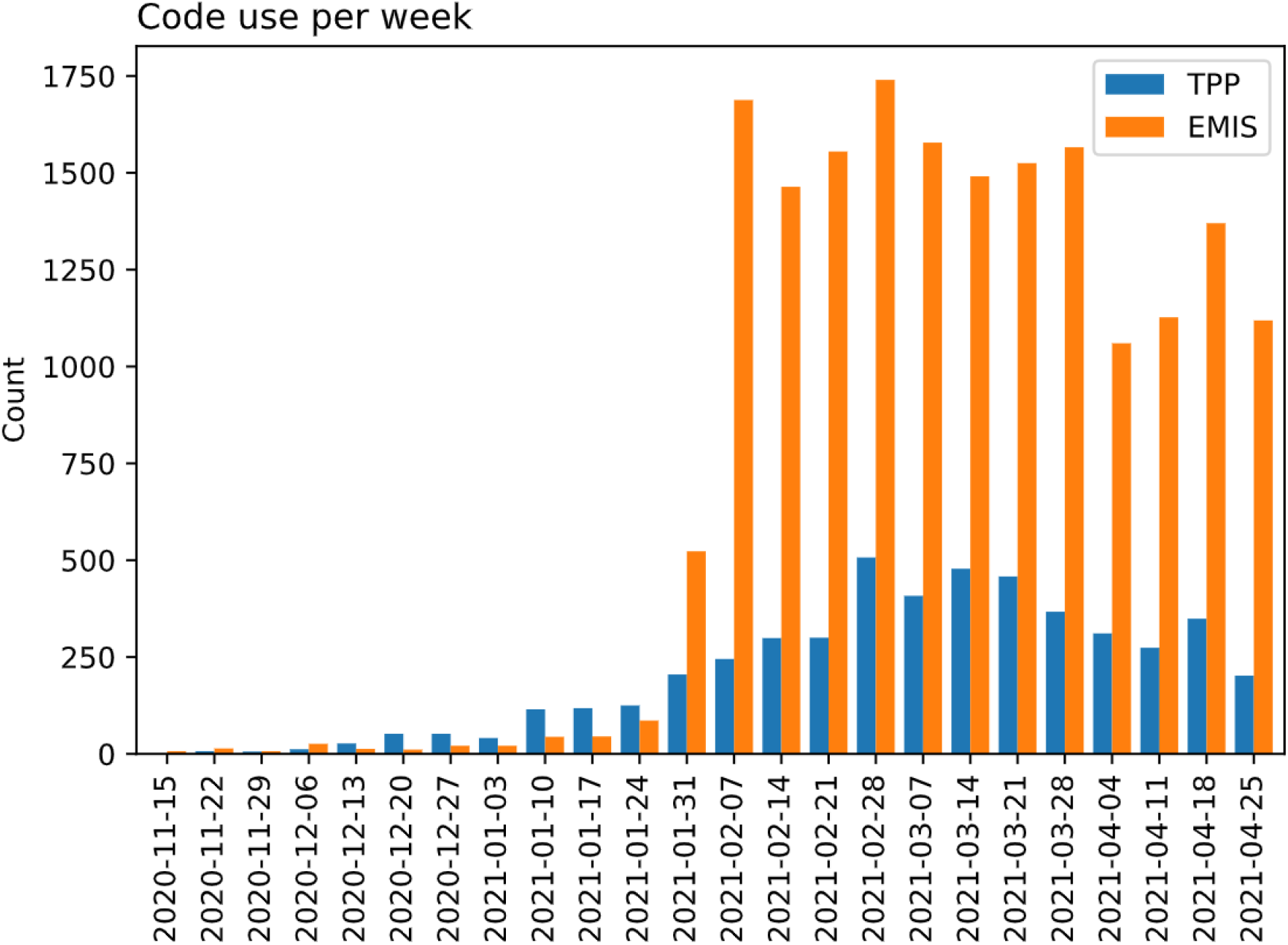
Use of long COVID codes over time, stratified by the electronic health record provider of the practice (TPP/SystmOne or EMIS). Reporting lag may affect recent dates

### Coding of individual SNOMED codes

The diagnostic codes were the most commonly used codes, particularly the “Post-COVID-19 syndrome” code, which accounted for 64.3% of all recorded codes. However there are differences in the distribution of codes between TPP and EMIS practices (Table 3). Codes relating to assessment of long COVID accounted for just 2.4% of long COVID codes used to date.

**Table 3:**
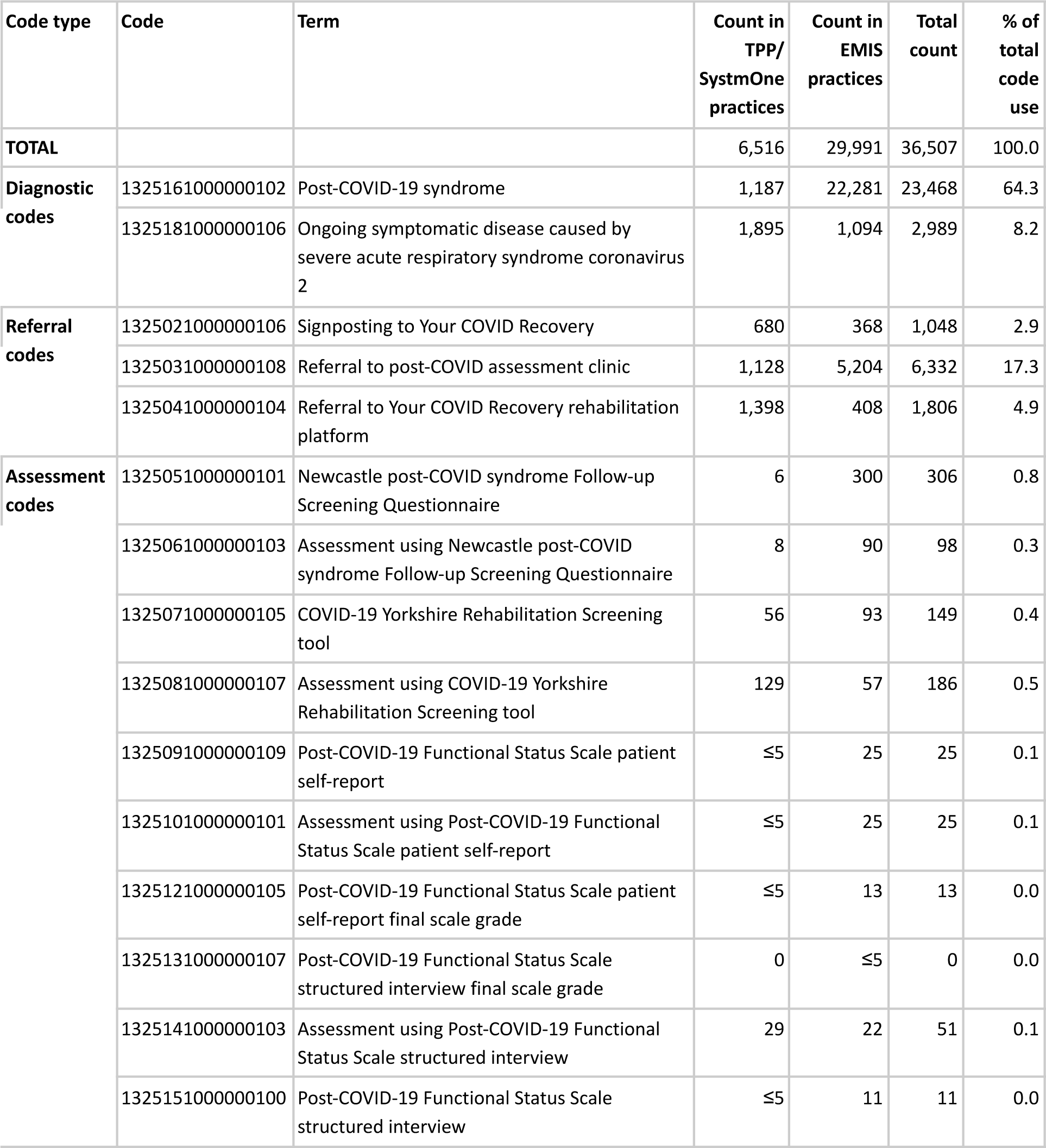
Total use of each individual long COVID related code. This is distinct from table 1 in that it counts all coded events, including where patients have been coded more than once.

| Code type | Code | Term | Count in TPP/<br>SystemOne practices | Count in EMIS practices | Total count | % of total code use |
| --- | --- | --- | --- | --- | --- | --- |
| **TOTAL** |  |  | 6,516 | 29,991 | 36,507 | 100.0 |
| **Diagnostic codes** | 1325161000000102 | Post-COVID-19 syndrome | 1,187 | 22,281 | 23,468 | 64.3 |
|  | 1325181000000106 | Ongoing symptomatic disease caused by severe acute respiratory syndrome coronavirus 2 | 1,895 | 1,094 | 2,989 | 8.2 |
| **Referral codes** | 1325021000000106 | Signposting to Your COVID Recovery | 680 | 368 | 1,048 | 2.9 |
|  | 1325031000000108 | Referral to post-COVID assessment clinic | 1,128 | 5,204 | 6,332 | 17.3 |
|  | 1325041000000104 | Referral to Your COVID Recovery rehabilitation platform | 1,398 | 408 | 1,806 | 4.9 |
| **Assessment codes** | 1325051000000101 | Newcastle post-COVID syndrome Follow-up Screening Questionnaire | 6 | 300 | 306 | 0.8 |
|  | 1325061000000103 | Assessment using Newcastle post-COVID syndrome Follow-up Screening Questionnaire | 8 | 90 | 98 | 0.3 |
|  | 1325071000000105 | COVID-19 Yorkshire Rehabilitation Screening tool | 56 | 93 | 149 | 0.4 |
|  | 1325081000000107 | Assessment using COVID-19 Yorkshire Rehabilitation Screening tool | 129 | 57 | 186 | 0.5 |
|  | 1325091000000109 | Post-COVID-19 Functional Status Scale patient self-report | ≤5 | 25 | 25 | 0.1 |
|  | 1325101000000101 | Assessment using Post-COVID-19 Functional Status Scale patient self-report | ≤5 | 25 | 25 | 0.1 |
|  | 1325121000000105 | Post-COVID-19 Functional Status Scale patient self-report final scale grade | ≤5 | 13 | 13 | 0.0 |
|  | 1325131000000107 | Post-COVID-19 Functional Status Scale structured interview final scale grade | 0 | ≤5 | 0 | 0.0 |
|  | 1325141000000103 | Assessment using Post-COVID-19 Functional Status Scale structured interview | 29 | 22 | 51 | 0.1 |
|  | 1325151000000100 | Post-COVID-19 Functional Status Scale structured interview | ≤5 | 11 | 11 | 0.0 |

## Discussion

### Summary

As of late April 2021, 23,273 people had a record of at least one long COVID code in their primary care record. Use between different general practices varied greatly, and a large proportion (26.7%) have never used any long COVID code. We found substantially higher recording in practices that use EMIS software compared to those who use TPP software. Amongst those people who did have a recorded long COVID code, rates were highest in the working age population and more common in women.

### Strengths and weaknesses

The key strength of this study is its unprecedented scale: we include over 58 million people, 95% of the population in England. In contrast with many studies that use electronic health record data, we were also able to compare long COVID diagnostic codes between practices that use different software systems, and find a striking disparity: this has important implications for understanding whether clinicians are using the codes appropriately. A key weakness of this data for estimating true prevalence of long COVID in primary care, and factors associated with the condition, is that it relies on clinicians formally entering a diagnostic or referral code into the patient’s record: we note that this is a limitation of all electronic health record research for all clinical conditions and activity; however the emergence of a new diagnosis and recent launch of a new set of diagnostic codes may present new challenges in this regard.

### Research in Context

To our knowledge there are no other studies on prevalence of long COVID using clinicians’ diagnoses or electronic health records data. There are numerous studies using self-reported data from patients on the prevalence of continued symptoms following COVID-19, with estimates varying between 4.5% and 89%, largely due to highly variable case definitions^4^; individual symptoms characteristing long COVID have been reported as fatigue, headache, dyspnea and anosmia^5^. The ONS COVID Infection Survey estimates prevalence of self-diagnosed long COVID at 13.7%^6^. Separately numerous cohort studies have reported an increased risk of serious cardiovascular and metabolic outcomes following hospitalisation with COVID^7,8^, and there are various prospective studies such as PHOSP following up hospitalised patients over the next year^9^.

### Interpretation and implications

The prevalence of long COVID codes in primary care is extremely low when compared with early reports of long COVID prevalence. The large variation in apparent rate of long COVID between different geographic regions, practices and electronic health record systems strongly suggests that clinicians’ coding practice is inconsistent at present. This may reflect variation in awareness of new diagnostic codes that were only launched in November 2020, and only available in EMIS at the end of January 2021. In addition, the codes for long COVID and associated synonyms do not currently contain the term “long COVID”: this was an active choice by NICE after detailed consideration of terminology in their guidance, and by NHS Digital who manage SNOMED-UK codes^10^. This has resulted in a mismatch between formal clinical terminology and popular parlance among clinicians and patients.

In our view those managing SNOMED terminology for England should either update the long COVID codes to include the phrase “long COVID”, ideally in advance of the upcoming new SNOMED international release; or energetically disseminate their preferred new phrasing to all frontline clinicians, to ensure more appropriate use of these codes. Similarly NICE and other authoritative bodies giving guidance on long COVID should energetically communicate to clinicians the importance of correctly coding long COVID in patient records. It is a high national priority to estimate the prevalence of long COVID, identify its causes and consequences, and plan services appropriately. This cannot be done when clinical terminology is ambiguously and inconsistently used.

The variation in recording between users of different electronic health record software is also striking. After speaking with clinicians and both software vendors, the reasons for this disparity remain unclear, but are likely attributable to differences in user interface, which has previously been shown to influence clinicians’ treatment choices^11,12^. This should be addressed by interviewing GPs about their experiences with diagnosing and treating people with long COVID in each system.

Despite these issues around correct recording of clinicians’ diagnoses, there also remains a strong possibility that clinicians are not currently diagnosing their patients as having long COVID. The prevalence of long COVID in self-report in patient surveys is substantially higher than we have found here. To our mind this disparity can only be resolved by conducting prospective surveys with clinicians themselves, evaluating how many patients they have seen with a condition they would understand to be diagnosable as long COVID, perhaps complemented with qualitative research on the topic.

If we accept that the different rates of long COVID usage in each sub-group reflects the true comparative risk for each demographic then there are two key findings: firstly, the lower rate in older patients, despite their higher prevalence of severe acute COVID outcomes^13^, which may be affected by the competing risk of death in COVID-19 patients; and secondly, the higher rate of long COVID in women, despite the higher prevalence of severe acute COVID outcomes in men, which may be explained in part by differences in routine consultation rates between men and women.^14^

## Conclusions

Current recording of long COVID in primary care is variable, and low. This may reflect under-coding, sub-optimal design and communication of clinical terms, under-diagnosis, a true low prevalence of long COVID diagnosed by clinicians, or a combination of factors. We will update this analysis regularly with extended follow-up time.

## Data Availability

All data were linked, stored, and analysed securely within the OpenSAFELY platform. Detailed pseudonymised patient data are potentially re-identifiable and therefore not shared. We rapidly delivered the OpenSAFELY data analysis platform without previous funding to deliver timely analyses of urgent research questions in the context of the global COVID-19 health emergency: now that the platform is established, we have established a process for external users to request access to the platform https://www.opensafely.org/onboarding-new-users/.

## Acknowledgements

We are very grateful for all the support received from the EMIS and TPP Technical Operations team throughout this work, and for generous assistance from the information governance and database teams at NHS England / NHSX.

## Information governance and ethical approval

NHS England is the data controller; EMIS and TPP are the data processors; and the key researchers on OpenSAFELY are acting on behalf of NHS England. This implementation of OpenSAFELY is hosted within the EMIS and TPP environments which are accredited to the ISO 27001 information security standard and are NHS IG Toolkit compliant;^15,16^ patient data has been pseudonymised for analysis and linkage using industry standard cryptographic hashing techniques; all pseudonymised datasets transmitted for linkage onto OpenSAFELY are encrypted; access to the platform is via a virtual private network (VPN) connection, restricted to a small group of researchers; the researchers hold contracts with NHS England and only access the platform to initiate database queries and statistical models; all database activity is logged; only aggregate statistical outputs leave the platform environment following best practice for anonymisation of results such as statistical disclosure control for low cell counts.^17^ The OpenSAFELY research platform adheres to the obligations of the UK General Data Protection Regulation (GDPR) and the Data Protection Act 2018. In March 2020, the Secretary of State for Health and Social Care used powers under the UK Health Service (Control of Patient Information) Regulations 2002 (COPI) to require organisations to process confidential patient information for the purposes of protecting public health, providing healthcare services to the public and monitoring and managing the COVID-19 outbreak and incidents of exposure; this sets aside the requirement for patient consent.^18^ Taken together, these provide the legal bases to link patient datasets on the OpenSAFELY platform. GP practices, from which the primary care data are obtained, are required to share relevant health information to support the public health response to the pandemic, and have been informed of the OpenSAFELY analytics platform. This study was approved by the Health Research Authority (REC reference 20/LO/0651) and by the LSHTM Ethics Board (reference 21863).

## Guarantor

BG/LS are guarantors of the OpenSAFELY project.

## Appendix

### Information governance and ethics

NHS England is the data controller; TPP is the data processor; and the key researchers on OpenSAFELY are acting on behalf of NHS England. OpenSAFELY is hosted within the TPP environment which is accredited to the ISO 27001 information security standard and is NHS IG Toolkit compliant;^19,20^ patient data are pseudonymised for analysis and linkage using industry standard cryptographic hashing techniques; all pseudonymised datasets transmitted for linkage onto OpenSAFELY are encrypted; access to the platform is via a virtual private network (VPN) connection, restricted to a small group of researchers who hold contracts with NHS England and only access the platform to initiate database queries and statistical models. Pseudonymised structured data include demographics, medications prescribed from primary care, diagnoses, and laboratory measures. No free text data are included. All database activity is logged; only aggregate statistical outputs leave the platform environment following best practice for anonymisation of results such as statistical disclosure control for low cell counts.^21^ The OpenSAFELY research platform adheres to the obligations of the UK General Data Protection Regulation (GDPR) and the Data Protection Act 2018. In March 2020, the Secretary of State for Health and Social Care used powers under the UK Health Service (Control of Patient Information) Regulations 2002 (COPI) to require organisations to process confidential patient information for the purposes of protecting public health, providing healthcare services to the public and monitoring and managing the COVID-19 outbreak and incidents of exposure; this sets aside the requirement for patient consent.^22^ Taken together, these provide the legal bases to link patient datasets on the OpenSAFELY platform. GP practices, from which the primary care data are obtained, are required to share relevant health information to support the public health response to the pandemic, and have been informed of the OpenSAFELY analytics platform. This study was approved by the Health Research Authority (REC reference 20/LO/0651) and by the LSHTM Ethics Board (ref 21863).

